# Genotype-first analysis in an unselected health system-based population reveals variable phenotypic severity of *COL4A5* variants

**DOI:** 10.1101/2024.06.04.24308453

**Authors:** McKenzie Zellers, Kaushal Solanki, Melissa A Kelly, Karyn M Murphy, Kyle Retterer, H. Les Kirchner, Ion Dan Bucaloiu, Bryn Moore, Tooraj Mirshahi, Alexander R Chang

**Author notes:** Corresponding author: Alexander R. Chang, MD, MS Center for Kidney Health Research Departments of Nephrology and Population Health Sciences Geisinger Medical Center 100 North Academy Avenue, Danville, PA 17822.

## Abstract

**Introduction:** Our knowledge of X-linked Alport Syndrome [AS] comes mostly from selected cohorts with more severe disease.

**Methods:** We examined the phenotypic spectrum of X-linked AS in males and females with a genotype-based approach using data from the Geisinger MyCode DiscovEHR study, an unselected health system-based cohort with exome sequencing and electronic health records. Patients with *COL4A5* variants reported as pathogenic (P) or likely pathogenic (LP) in ClinVar, or protein-truncating variants (PTVs), were each matched with up to 5 controls without *COL4A3/4/5* variants by sociodemographics, diabetes diagnosis, and year of first outpatient encounter. AS-related phenotypes included dipstick hematuria, bilateral sensorineural hearing loss (BSHL), proteinuria, decreased eGFR, and ESKD.

**Results:** Out of 170,856 patients, there were 29 hemizygous males (mean age 52.0 y [SD 20.0]) and 55 heterozygous females (mean age 59.3 y [SD 18.8]) with a *COL4A5* P/LP variant, including 48 with the hypomorphic variant p.Gly624Asp. Overall, penetrance (having any AS phenotypic feature) was highest for non-p.Gly624Asp P/LP variants (males: 94%, females: 85%), intermediate for p.Gly624Asp (males: 77%, females: 69%), compared to controls (males: 32%; females: 50%). The proportion with ESKD was highest for males with P/LP variants (44%), intermediate for males with p.Gly624Asp (15%) and females with P/LP variants (10%), compared to controls (males: 3%, females 2%). Only 47% of individuals with *COL4A5* had completed albuminuria screening, and a minority were taking renin-angiotensin aldosterone system (RAAS) inhibitors. Only 38% of males and 16% of females had a known diagnosis of Alport syndrome or thin basement membrane disease.

**Conclusion:** In an unselected cohort, we show increased risks of AS-related phenotypes in men and women compared to matched controls, while showing a wider spectrum of severity than has been described previously and variability by genotype. Future studies are needed to determine whether early genetic diagnosis can improve outcomes in Alport Syndrome.

## Introduction

Alport syndrome (AS) is the second most common genetic cause of end stage kidney disease behind polycystic kidney disease, affecting approximately one in 2320 individuals^1,2^. Nearly 85% of total cases of AS in the past have been due to variants in the *COL4A5* gene found on the X chromosome, with the rest of individuals with AS having variants in the *COL4A3* and *COL4A4* genes responsible for autosomal recessive and dominant inheritance patterns^1^. Individuals with AS can have phenotypic manifestations related to abnormalities in the basement membranes of the glomerulus, cochlea of the ear, and lens of the eye, resulting in microscopic hematuria, sensorineural hearing loss, and ocular disturbances. Each collagenous IV chain contains a collagenous domain made of Gly Xaa Yaa triplet repeats, resulting in a rigid protein structure^3^. Thus, glycine missense variants disrupting the intermediate collagenous domains as well as protein-truncating variants (PTVs) are likely to be deleterious. Some but not all studies suggest a worse prognosis for PTVs than missense variants^4–8^. One glycine missense variant, p.Gly624Asp, is adjacent to a non-collagenous region and known to have a more mild phenotype.^9^

Although females are more likely to carry *COL4A5* variants due to the nature of the X-linked inheritance pattern, males are more severely impacted, with reported risk of progression to ESKD reported at nearly 100% in *COL4A5* males compared to 25% in similarly affected females^10,11^. An important limitation to the existing literature is that participants in these studies have been selected from families known to be affected by AS, likely leading to an overestimation of penetrance and phenotypic severity. As availability of genetic testing increases and costs of testing decrease, there is an urgent need to understand the penetrance and phenotypic spectrum in the general population to inform families about the implications of a genetic diagnosis of AS.

Using a genotype-first approach, we aimed to examine the phenotypic spectrum of X-linked AS due to *COL4A5* variants using an unselected health system-based population. We hypothesized that both males and females with *COL4A5* variants would have higher risk of kidney phenotypes though the penetrance and phenotypic severity of disease would be lower than previously reported in more selected cohorts.

## Methods

### Study Population

There were 170,856 research participants in MyCode who had whole exome sequencing data available after quality control. These research participants are patients in the Geisinger Health System at large who consented to participate in the MyCode Community Health Initiative and were sequenced through the Geisinger-Regeneron DiscovEHR collaboration^12^. This research study was approved by the Geisinger Institutional Review Board. Patients with ClinVar P/LP or PTVs, as defined below, were classified as cases. We queried ClinVar [date accessed 4/24/2023] for *COL4A5* (NM_033380.3) variants previously classified as pathogenic (P) or likely pathogenic (LP) by >1 submitter and no conflicts, or conflicting variants with a ratio of ≥6 P/LP to other classifications^13^. Predicted protein truncating variants (PTVs; frameshift, nonsense, splice) were identified from those called as high impact variants by variant effect predictor v109. All variants underwent manual review to ensure technical confidence and clinical relevance.^14^ Any variants that did not meet quality thresholds were confirmed in a Clinical Laboratory Improvement Amendments (CLIA)-certified laboratory using orthogonal technology prior to inclusion.

### Phenotypic outcomes

Phenotypic data sources included International Classification of Diseases (ICD) 9 and 10 codes, blood and urine laboratory data, and chart review. ICD codes were used for hypertension, diabetes, hearing loss, bilateral sensorineural hearing loss, AS, focal segmental glomerulosclerosis (FSGS), and ESKD (**Supplemental Table 1**). We used the 2021 CKD-EPI equation to calculate eGFR, and we imputed eGFR of 5 ml/min/1.73m^2^ for patients with kidney transplant.^15^ Hematuria on dipstick was defined as having trace blood or greater on at least half of all urinalyses, excluding urinalyses positive for leukocyte esterase or nitrites.^5^ Age at time of ESKD was determined using linkage to the United States Renal Data System,^16^ and if these data were not available, by chart review. Presence of albuminuria was defined as ever having albumin to creatinine ratio (ACR) ≥ 30 mg/g, and presence of proteinuria by dipstick was defined requiring at least 2 dipsticks. Any phenotypic feature of AS was defined as hematuria on urinalysis or ICD code, albuminuria, eGFR <60 ml/min/1.73m^2^, FSGS ICD code, ESKD ICD code, or bilateral sensorineural hearing loss. Patients with most recent eGFR data at age ≥18 years were categorized into Kidney Disease Improving Global Outcomes (KDIGO) risk categories using estimated glomerular filtration rate (eGFR) and ACR (**Supplemental Table 2**).^5,17^ If ACR was not available, urine dipstick protein data was used for KDIGO risk categorization with dipstick protein 1+ twice classified as ACR 30-299 mg/g, and dipstick protein 2+ or greater twice classified as ACR 300+ mg/g. Chart review was performed on all cases to further examine diagnosis of AS or TBMD, kidney biopsies, treatment with angiotensin-converting enzyme inhibitors (ACEIs) or angiotensin receptor blockers (ARBs), family history, and genetic testing.

### Statistical analysis

Patients with *COL4A5* P/LP variants were matched to up to 5 controls (patients without *COL4A3, COL4A4,* and *COL4A5* variants at allele frequency<0.01), with exact matching on age (within 1 year) at time of most recent eGFR, sex, race, diabetes diagnosis, and year of first outpatient encounter. We compared characteristics between patients with *COL4A5* P/LP variants and matched controls using chi-square tests for categorical outcomes and paired t-tests for continuous outcomes overall, and by sex and variant group. We used Poisson regression models with robust variance as the primary analysis to estimate prevalence ratios to determine AS-related phenotypic outcomes, stratified by sex and genotype (P/LP variant or the known hypomorphic p.Gly624Asp variant), using each subgroups’ matched controls. ^18,19^ We also conducted time-to-event analyses with ESKD as the outcome using Kaplan-Meier survival curves with age as the time variable. Patients were censored at age of last eGFR value. The proportional hazards assumption was graphically assessed, and Cox proportional hazards models were adjusted for age at time of first eGFR value in sex-stratified analyses. As prior *COL4A5* studies have suggested genotype-phenotype correlations with the worst outcomes for nonsense and frameshift variants, intermediate outcomes for splice variants, and milder phenotypes for missense and in-frame deletion variants,^6,20,21^ we also estimated risk of ESKD using Cox models for nonsense/frameshift variants, splice variants, the hypomorphic p.Gly624Asp variant, and other missense or in-frame deletion variants. We conducted sensitivity analyses, including time-to-event competing-risks regression using the method of Fine and Gray,^22^ and analyses using Firth logistic regression, including only one individual from each family. We considered p values <0.05 to be significant. Analyses were conducted using STATA/MP 15.1.

## Results

Out of the 170,856 patients, there were 29 hemizygous males (mean age 52.0 y [SD 20.0]) and 55 heterozygous females (mean age 59.3 y [SD 18.8]) with a *COL4A5* P/LP variant. The most common *COL4A5* P/LP variant was p.Gly624Asp (n=48), and there were 17 other unique variants, including 4 other missense or in-frame deletion variants (n=16 individuals), 7 splicing variants (n=13 individuals), 4 frameshift variants (n=4 individuals), and 2 stop-gained variants (n=3 individuals). Of the 84 individuals with *COL4A5* P/LP variants, the vast majority were matched to 5 controls (78 individuals matched to 5 controls, 2 to 3 controls, 2 to 2 controls, 2 to 1 control). Compared to matched controls, individuals with *COL4A5* P/LP variants were more likely to have urinalysis (96% vs 82%, p=0.001) and ACR (47% vs 26%, p<0.001) results available in addition to having a hypertension diagnosis (68% vs 51%, p=0.004) whereas 100% of patients in both groups had evaluation of eGFR (**Table 1**). The median (interquartile interval) year of first outpatient visit was 2003.5 (2001-2009.5) for individuals with *COL4A5* and 2004 (2001-2009) for matched controls.

**Table 1.**
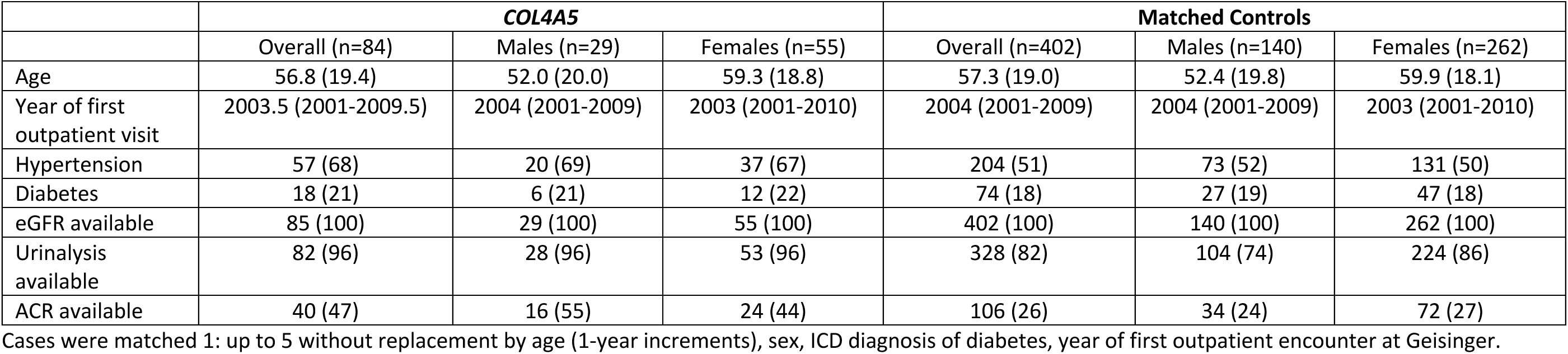
Characteristics of *COL4A5* P/LP X-linked Alport Syndrome patients and matched controls.

### Prevalence of AS Phenotypes by Sex

Overall, males with *COL4A5* P/LP variants had the highest risk of AS-related phenotypes, followed by males with the hypomorphic p.Gly624Asp variant and females with *COL4A5* P/LP variants, with lowest risk in females with p.Gly624Asp (**Table 2**, **Figure 1**). The p.Gly624Asp variant was associated with a milder phenotype than other P/LP variants for both males and females. Males with p.Gly624Asp demonstrated lower penetrance (n=13; 0% AS ICD, 15% ESKD ICD, 31% hearing loss ICD, 23% eGFR <30 ml/min/1.73m^2^) compared to other hemizygous *COL4A5* P/LP males (n=16; 31% AS ICD, 44% ESKD ICD, 50% hearing loss ICD, 44% eGFR <30 ml/min/1.73m^2^). KDIGO risk categorization demonstrated that 33% of p.Gly624Asp males (mean age 55.7 y [SD 17.1] had a high-risk category or greater as compared to 80% of other hemizygous *COL4A5* P/LP males (mean age 49.0 y [SD 22.2]) (**Table 2**, **Figure 1**). Similarly, the prevalence of high-risk KDIGO category or greater in p.Gly624Asp females (mean age 60.3 y [SD 14.8] was 24% compared to 68% of other P/LP females (mean age 59.9 y [SD 18.1]).

**Figure 1.**
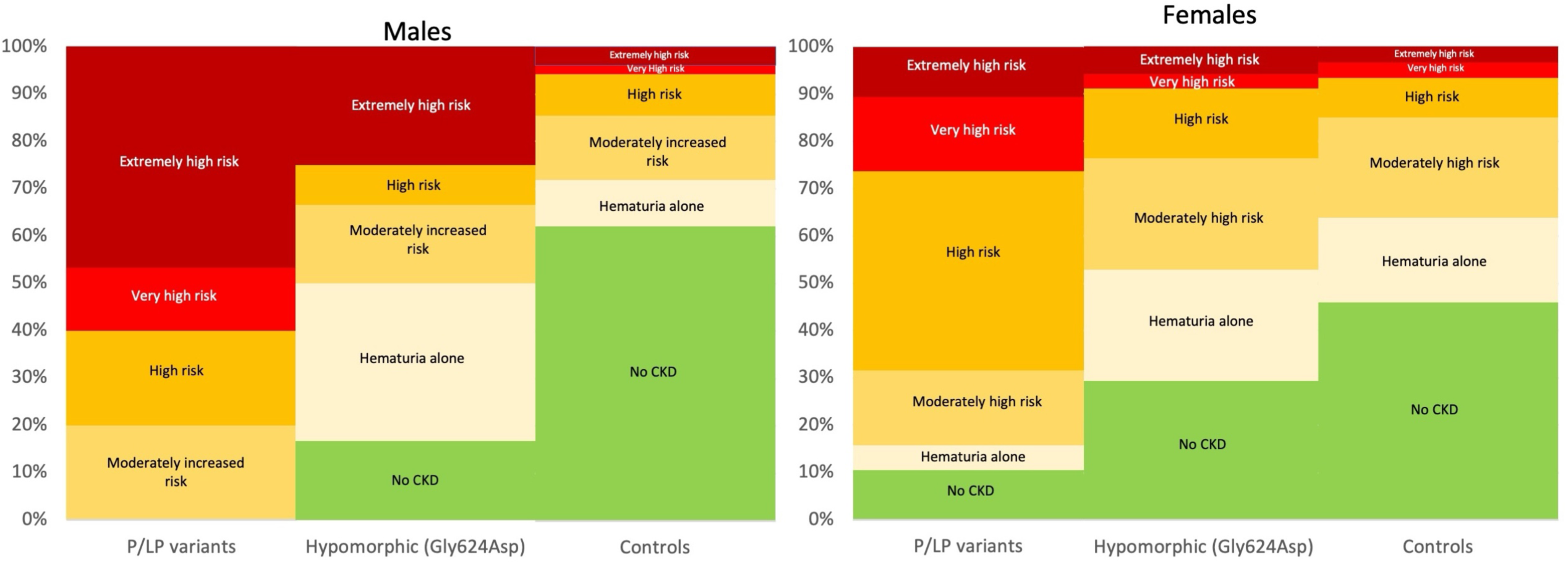
KDIGO risk categories by sex and variant type. KDIGO risk categories created using eGFR and ACR. If ACR not available, then urine dipstick protein data used with dipstick protein 1+ twice classified as ACR 30-299 mg/g, and dipstick protein 2+ or greater twice classified as ACR 300+ mg/g. KDIGO risk categories defined as: 0 - no CKD or hematuria; 1 - hematuria alone (eGFR ≥60, ACR <30); 2 – Moderately increased risk (eGFR 45-59 and ACR<30 or eGFR ≥60 with ACR 30-299); 3 – High risk (eGFR 30-44 with ACR<30 or eGFR 45-59 with ACR 30-299, or eGFR ≥60 with ACR 300+); 4 – Very high risk (eGFR 15-29 with ACR<300 or eGFR 30-44 with ACR ≥30 or eGFR 45-59 with ACR ≥300); 5 – Extremely high risk (eGFR<15 or eGFR 15-29 with ACR 300+; see Supplemental Table 2). Two cases and 6 controls <18 years of age were excluded from this categorization.

**Table 2.**
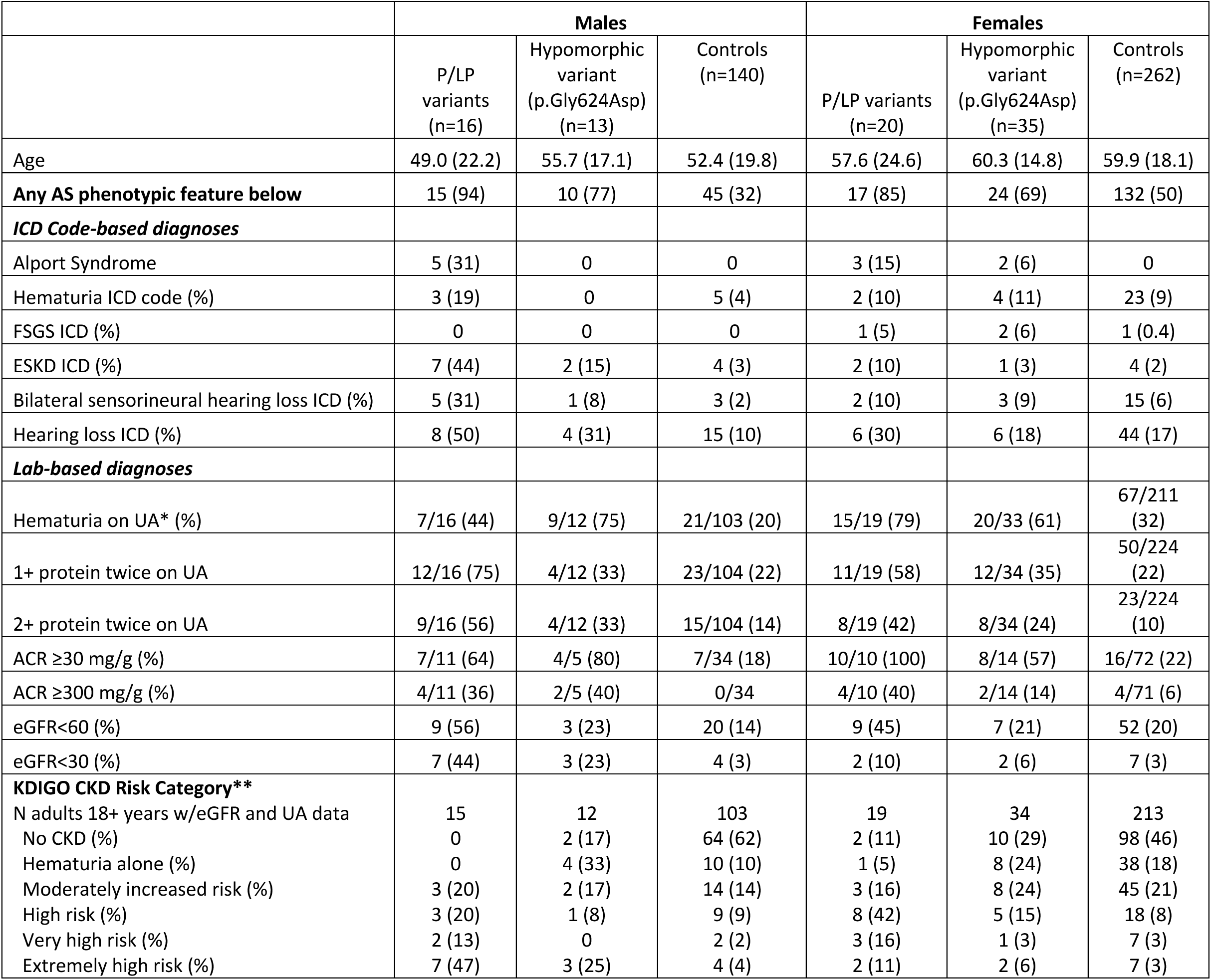
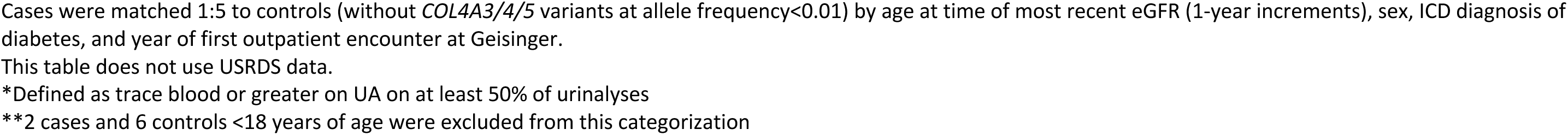
Phenotypic Features of *COL4A5* P/LP X-linked Alport Syndrome by Sex and Genotype.

|  | Males |  |  | Females |  |  |
| --- | --- | --- | --- | --- | --- | --- |
|  | P/LP variants (n=16) | Hypomorphic variant (p.Gly624Asp) (n=13) | Controls (n=140) | P/LP variants (n=20) | Hypomorphic variant (p.Gly624Asp) (n=35) | Controls (n=262) |
| Age | 49.0 (22.2) | 55.7 (17.1) | 52.4 (19.8) | 57.6 (24.6) | 60.3 (14.8) | 59.9 (18.1) |
| **Any AS phenotypic feature below** | 15 (94) | 10 (77) | 45 (32) | 17 (85) | 24 (69) | 132 (50) |
| **ICD Code-based diagnoses** |  |  |  |  |  |  |
| Alport Syndrome | 5 (31) | 0 | 0 | 3 (15) | 2 (6) | 0 |
| Hematuria ICD code (%) | 3 (19) | 0 | 5 (4) | 2 (10) | 4 (11) | 23 (9) |
| FSGS ICD (%) | 0 | 0 | 0 | 1 (5) | 2 (6) | 1 (0.4) |
| ESKD ICD (%) | 7 (44) | 2 (15) | 4 (3) | 2 (10) | 1 (3) | 4 (2) |
| Bilateral sensorineural hearing loss ICD (%) | 5 (31) | 1 (8) | 3 (2) | 2 (10) | 3 (9) | 15 (6) |
| Hearing loss ICD (%) | 8 (50) | 4 (31) | 15 (10) | 6 (30) | 6 (18) | 44 (17) |
| **Lab-based diagnoses** |  |  |  |  |  |  |
| Hematuria on UA* (%) | 7/16 (44) | 9/12 (75) | 21/103 (20) | 15/19 (79) | 20/33 (61) | 67/211 (32) |
| 1+ protein twice on UA | 12/16 (75) | 4/12 (33) | 23/104 (22) | 11/19 (58) | 12/34 (35) | 50/224 (22) |
| 2+ protein twice on UA | 9/16 (56) | 4/12 (33) | 15/104 (14) | 8/19 (42) | 8/34 (24) | 23/224 (10) |
| ACR ≥30 mg/g (%) | 7/11 (64) | 4/5 (80) | 7/34 (18) | 10/10 (100) | 8/14 (57) | 16/72 (22) |
| ACR ≥300 mg/g (%) | 4/11 (36) | 2/5 (40) | 0/34 | 4/10 (40) | 2/14 (14) | 4/71 (6) |
| eGFR<60 (%) | 9 (56) | 3 (23) | 20 (14) | 9 (45) | 7 (21) | 52 (20) |
| eGFR<30 (%) | 7 (44) | 3 (23) | 4 (3) | 2 (10) | 2 (6) | 7 (3) |
| **KDIGO CKD Risk Category\*\*** |  |  |  |  |  |  |
| N adults 18+ years w/eGFR and UA data | 15 | 12 | 103 | 19 | 34 | 213 |
| No CKD (%) | 0 | 2 (17) | 64 (62) | 2 (11) | 10 (29) | 98 (46) |
| Hematuria alone (%) | 0 | 4 (33) | 10 (10) | 1 (5) | 8 (24) | 38 (18) |
| Moderately increased risk (%) | 3 (20) | 2 (17) | 14 (14) | 3 (16) | 8 (24) | 45 (21) |
| High risk (%) | 3 (20) | 1 (8) | 9 (9) | 8 (42) | 5 (15) | 18 (8) |
| Very high risk (%) | 2 (13) | 0 | 2 (2) | 3 (16) | 1 (3) | 7 (3) |
| Extremely high risk (%) | 7 (47) | 3 (25) | 4 (4) | 2 (11) | 2 (6) | 7 (3) |

Cases were matched 1:5 to controls (without *COL4A3/4/5* variants at allele frequency <0.01) by age at time of most recent eGFR (1-year increments), sex, ICD diagnosis of diabetes, and year of first outpatient encounter at Geisinger.
This table does not use USRDS data.
\*Defined as trace blood or greater on UA on at least 50% of urinalyses
\*\*2 cases and 6 controls <18 years of age were excluded from this categorization

All groups had significantly higher risk of AS phenotypic features compared to matched controls with prevalence ratios (PR) of 3.13 (95% CI: 2.18, 4.48) for males with P/LP variants, 2.20 (95% CI: 1.39, 3.48) for males with p.Gly624Asp, 1.75 (95% CI: 1.34, 2.29) for females with P/LP variants, and 1.35 (95% CI: 1.03, 1.76) for females with p.Gly624Asp (**Table 3**, **Figure 2**). Risks of the most common AS manifestation, dipstick hematuria, were similar for males with P/LP variants (PR 2.36, 95% CI: 1.07, 5.23), males with p.Gly624Asp (PR 3.34, 95% CI: 1.80, 6.21), females with P/LP variants (PR 2.59, 95% CI: 1.73, 3.87), and females with p.Gly624Asp (PR 1.86, 95% CI: 1.28, 2.70). Males and females with p.Gly624Asp tended to have milder phenotypic presentation than their counterparts with other P/LP variants. For example, prevalence ratios for ESKD were 35.00 (95% CI: 4.57, 268.10) for males with P/LP variants, 3.08 (95% CI: 0.56, 16.80) for males with p.Gly624Asp, 9.30 (95% CI: 0.88, 98.67) for females with P/LP variants, and 1.61 (95% CI: 0.17, 15.07) for females with p.Gly624Asp. Males with P/LP and p.Gly624Asp variants had significantly increased risk of hearing loss ICD diagnosis whereas females with P/LP variants or p.Gly624Asp did not. Sensitivity analyses restricted to one individual per family showed consistent results (**Supplemental Tables 3**). Variant-level phenotypic characteristics, stratified by sex, are shown in **Supplemental Table 4**.

**Figure 2.**
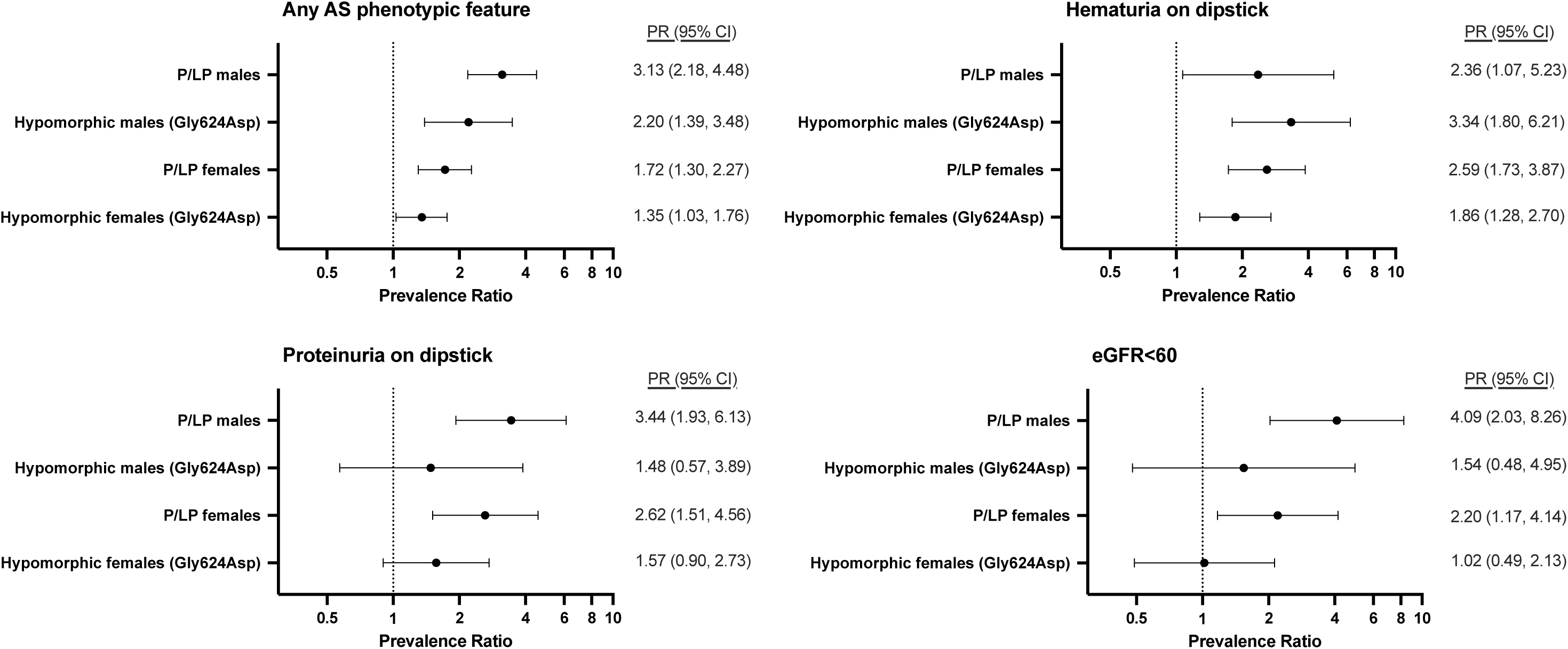
Prevalence Ratios for Alport Syndrome-related phenotypes. Associations between *COL4A5* P/LP variants and renal phenotypes examined using Poisson regression with robust variance in analyses in stratified analyses by genotype and sex.

**Table 3.** Prevalence Ratios of Alport Syndrome Phenotypic Features by Sex and Genotype.

|  | Males |  | Females |  |
| --- | --- | --- | --- | --- |
|  | P/LP variants (other than p.Gly624Asp)<br>Prevalence Ratio (95%) | p.Gly624Asp<br>Prevalence Ratio (95%) | P/LP variants (other than p.Gly624Asp)<br>Prevalence Ratio (95%) | p.Gly624Asp<br>Prevalence Ratio (95%) |
| **Any AS phenotypic feature below** | 3.13 (2.18, 4.48)*** | 2.20 (1.39, 3.48)** | 1.72 (1.30, 2.27)*** | 1.35 (1.03, 1.76)* |
| *ICD diagnosis-based* |  |  |  |  |
| **ESKD ICD** | 35.00 (4.57, 268.10)** | 3.08 (0.56, 16.80) | 9.30 (0.88, 98.67) | 1.61 (0.17, 15.07) |
| **Bilateral sensorineural hearing loss ICD** | 12.50 (2.63, 59.34)** | 4.62 (0.30, 70.41) | 1.03 (0.24, 4.45) | 2.41 (0.63, 9.23) |
| **Hearing loss ICD** | 3.08 (1.53, 6.21)** | 9.23 (1.87, 45.67)** | 1.16 (0.55, 2.48) | 1.49 (0.65, 3.44) |
| *Lab-based diagnoses* |  |  |  |  |
| **Trace blood or greater on UA\*** | 2.36 (1.07, 5.23)* | 3.34 (1.80, 6.21)*** | 2.59 (1.73, 3.87)*** | 1.86 (1.28, 2.70)** |
| **1+ protein twice on UA** | 3.44 (1.93, 6.13)*** | 1.48 (0.57, 3.89) | 2.62 (1.51, 4.56)** | 1.57 (0.90, 2.73) |
| **2+ protein twice on UA** | 4.42 (1.94, 10.05)*** | 2.04 (0.73, 5.71) | 3.62 (1.64, 7.98)** | 2.50 (1.12, 5.55)* |
| **ACR ≥30 mg/g** | 2.86 (1.06, 7.71)* | 6.40 (1.57, 26.01)** | 5.80 (2.59, 13.01)*** | 2.23 (1.12, 4.45)* |
| **eGFR<60** | 4.09 (2.03, 8.26)*** | 1.54 (0.48, 4.95) | 2.20 (1.17, 4.14)* | 1.02 (0.49, 2.13) |
| **eGFR<30** | 35.00 (4.57, 268.10)** | 4.62 (1.04, 20.56)* | 4.40 (0.65, 29.64) | 1.87 (0.38, 9.31) |

Each patient with *COL4A5* P/LP variant matched with up to 5 controls (without *COL4A3/4/5* variants at allele frequency <0.01). Poisson regression analyses with robust variance performed in sex-stratified analyses. For kidney transplant patients, eGFR imputed as 5 ml/min/1.73m<sup>2</sup>
This table does not use USRDS data.
\*P<0.05, \*\*P<0.01, \*\*\*P<0.001

### Time-to-event Analyses for ESKD by Genotype

Kaplan-Meier survival curves for ESKD-free survival followed similar patterns (**Figure 3**). Overall, there was no difference in deaths in patients with *COL4A5* variants and controls (7% vs. 7%; p=0.95) with mean age of death of *COL4A5* patients 70.8 y (range 54.3-80.7). **Table 4** shows hazard ratios (HR) for ESKD were significant for males with P/LP variants (HR 20.14, 95% CI: 5.85, 69.38; p<0.001), females with P/LP variants (HR 5.65, 95% CI: 1.00, 31.75), and of borderline significance for males with the hypomorphic p.Gly624Asp variant (HR 5.38, 95% CI: 0.97, 29.75). Females with the hypomorphic p.Gly624Asp variant were not at significantly increased risk of ESKD (HR 1.95, 95% CI: 0.22, 17.59). When examined by additional variant types, (e.g. nonsense/frameshift variants, splice, hypomorphic p.Gly624Asp, other missense or in-frame), risk was similar between variant groups though 95% intervals were very wide due to small sample sizes (**Supplemental Table 5)**. Results accounting for competing risks of death were consistent (**Supplemental Table 6)**. Mean age of ESKD onset in males was 36.9 y (SD 15.0) males with P/LP variants and 50.0 (SD 5.7) for males with Gly624Asp. Mean age of ESKD onset was 51.7 (SD 12.6) for females with no difference between those with P/LP or Gly624Asp variants.

**Figure 3.**
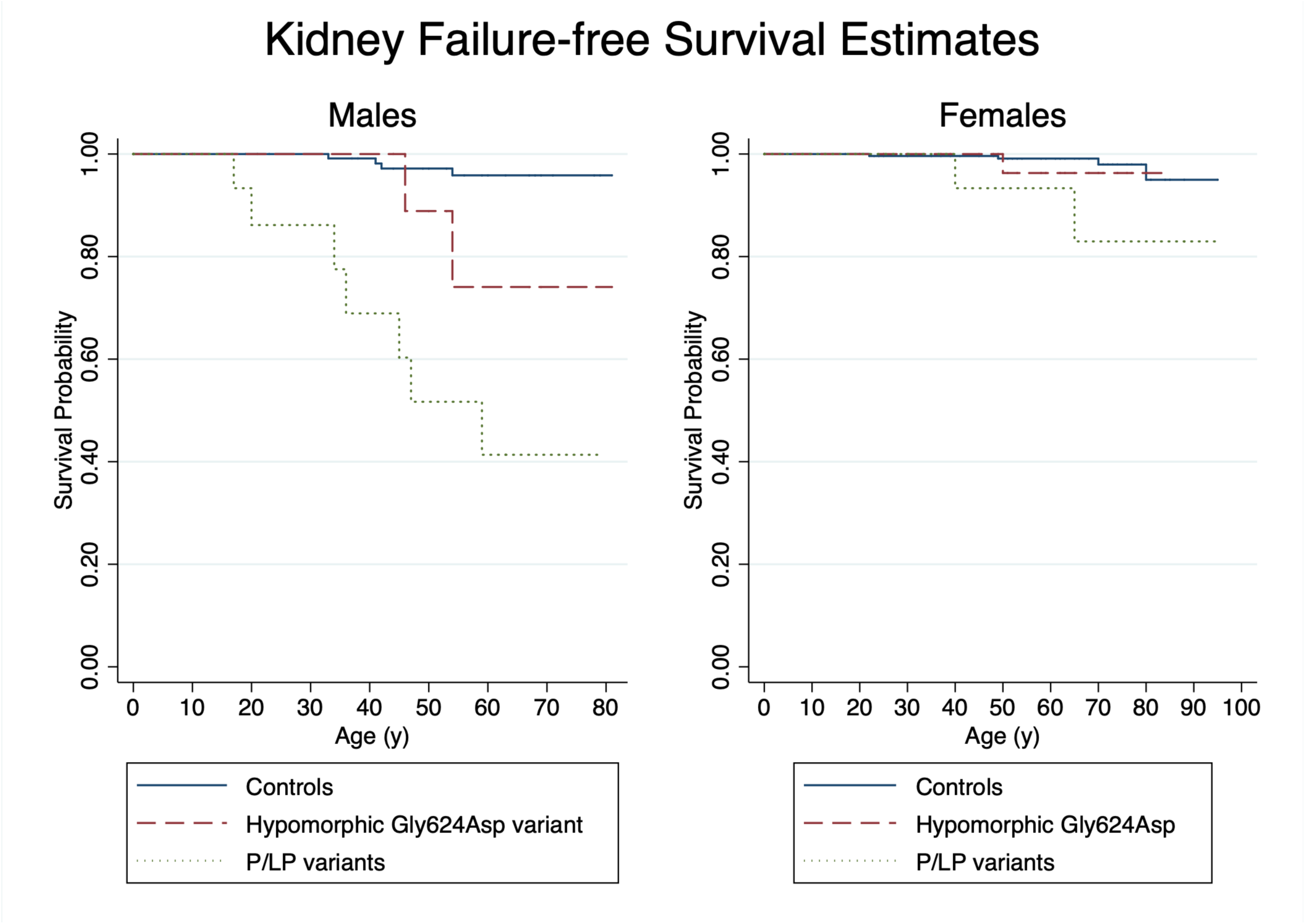
Kaplan-Meier Survival Curves for ESKD-free Survival. Kaplan-Meier curves comparing risk of ESKD by genotype for females and males with age as the time variable.

**Table 4.** Event Rates and Hazard Ratios for ESKD by Genotype in Males and Females.

|  | Event Rate (per 1000 person-years) | HR (95% CI) | P value |
| --- | --- | --- | --- |
| **Males** |  |  |  |
| Control (n=140) | 0.55 (0.20, 1.47) | Ref |  |
| Hypomorphic p.Gly624Asp (n=13) | 2.82 (0.71, 11.30) | 5.38 (0.97, 29.75) | 0.054 |
| P/LP variants (n=16) | 10.35 (4.94, 21.72) | 20.14 (5.85, 69.38) | <0.001 |
| **Females** |  |  |  |
| Control (n=262) | 0.26 (0.10, 0.68) | Ref |  |
| Hypomorphic p.Gly624Asp (n=35) | 0.48 (0.07, 3.39) | 1.95 (0.22, 17.59) | 0.553 |
| P/LP variants (n=20) | 1.80 (0.45, 7.21) | 5.65 (1.00, 31.75) | 0.049 |

Cox proportional hazards models adjusted for age (at time of first eGFR) in sex-stratified analyses.

### Individuals with a *COL4A5* P/LP variant and a second *COL4A3/4/5* variant

There were 15 individuals who had an additional rare (AF<0.01) variant in *COL4A3/4/5* with 4 (27%) having ESKD (**Supplemental Table 7)**. The individuals with ESKD included a male with *COL4A5* splice variant c.438+2T>G and *COL4A4* p.Pro352Leu; a male with *COL4A5* p.Leu1655Arg and *COL4A3* p.Val950Ile variant; a male with *COL4A5* p.Leu1655Arg and *COL4A3* p.Thr1489Ile, and a female with *COL4A5* c.4964T>G and *COL4A3* p.Ile1659Val.

### Diagnosis of AS

AS or thin basement membrane disease was documented in only 10/16 (63%) of males with P/LP variants, 1/13 (8%) of males with p.Gly624Asp, 4/20 (20%) of females with P/LP variants, and 4/35 (11%) of females with p.Gly624Asp. A total of 17 individuals (20%) had documentation of a native kidney biopsy performed, and none had undergone a skin biopsy to diagnose AS. Of those with native kidney biopsies, 9 (53%) had a biopsy diagnosis of AS, hereditary nephritis or thin basement membrane disease; FSGS was reported in 6 (33%) biopsies (**Table 5**). One patient had 2 biopsies as a child that were inconclusive (1^st^ biopsy: insufficient sample, thin basement membrane disease; 2^nd^ biopsy: IgM nephropathy, FSGS) and has yet to be diagnosed with AS. Only 2 (12%) had specific mention of staining for the alpha5 chain of type IV collagen. In terms of genetic diagnosis, only 5 (6%) received genetic testing and 2 (2%) additional patients reported having a family member who had genetic testing confirming diagnosis.

**Table 5.**
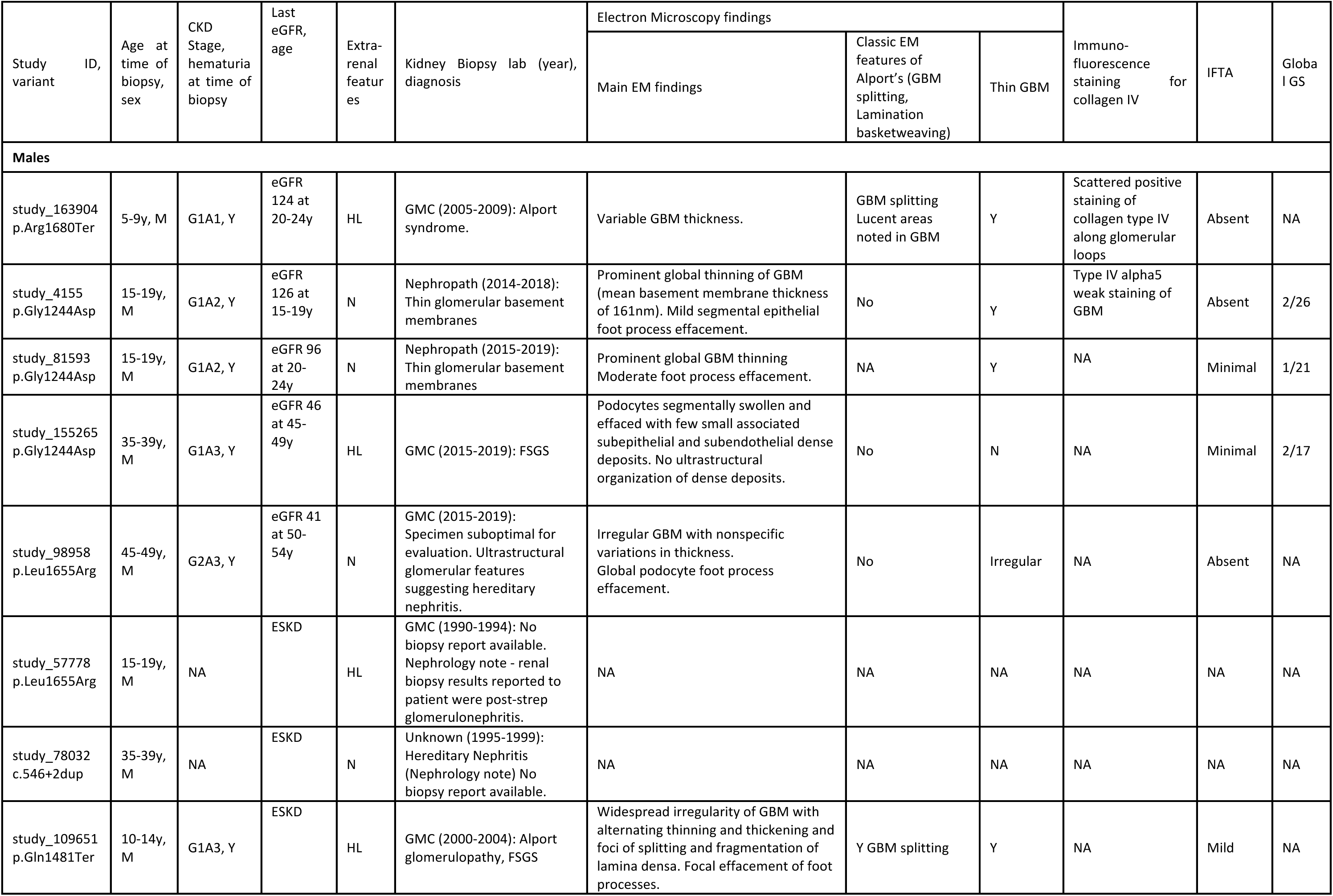
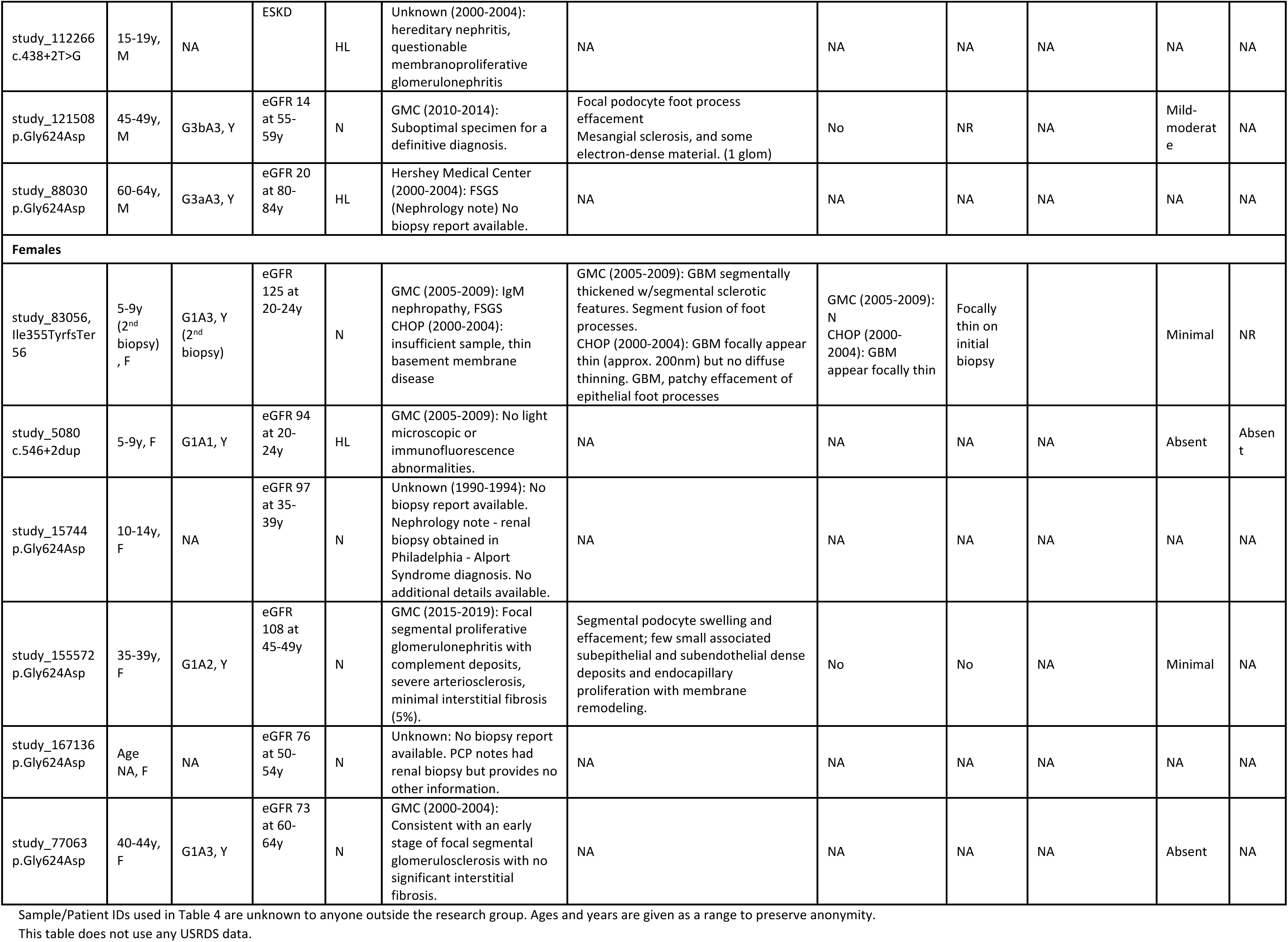
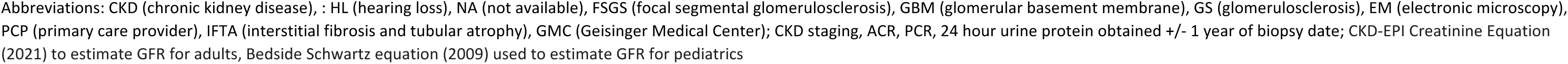
Characteristics of *COL4A5* P/LP X-linked Alport Syndrome with native kidney biopsies.

| Study variant ID, | Age at time of biopsy, sex | CKD Stage, hematuria at time of biopsy | Last eGFR, age | Extra-renal features | Kidney Biopsy lab (year), diagnosis | Electron Microscopy findings |  |  | Immuno-fluorescence staining for collagen IV | IFTA | Global GS |
| --- | --- | --- | --- | --- | --- | --- | --- | --- | --- | --- | --- |
|  |  |  |  |  |  | Main EM findings | Classic EM features of Alport's (GBM splitting, Lamination basketweaving) | Thin GBM |  |  |  |
| Males |  |  |  |  |  |  |  |  |  |  |  |
| study_163904 p.Arg1680Ter | 5-9y, M | G1A1, Y | eGFR 124 at 20-24y | HL | GMC (2005-2009): Alport syndrome. | Variable GBM thickness. | GBM splitting<br>Lucent areas noted in GBM | Y | Scattered positive staining of collagen type IV along glomerular loops | Absent | NA |
| study_4155 p.Gly1244Asp | 15-19y, M | G1A2, Y | eGFR 126 at 15-19y | N | Nephropath (2014-2018): Thin glomerular basement membranes | Prominent global thinning of GBM (mean basement membrane thickness of 161nm). Mild segmental epithelial foot process effacement. | No | Y | Type IV alpha5 weak staining of GBM | Absent | 2/26 |
| study_81593 p.Gly1244Asp | 15-19y, M | G1A2, Y | eGFR 96 at 20-24y | N | Nephropath (2015-2019): Thin glomerular basement membranes | Prominent global GBM thinning<br>Moderate foot process effacement. | NA | Y | NA | Minimal | 1/21 |
| study_155265 p.Gly1244Asp | 35-39y, M | G1A3, Y | eGFR 46 at 45-49y | HL | GMC (2015-2019): FSGS | Podocytes segmentally swollen and effaced with few small associated subepithelial and subendothelial dense deposits. No ultrastructural organization of dense deposits. | No | N | NA | Minimal | 2/17 |
| study_98958 p.Leu1655Arg | 45-49y, M | G2A3, Y | eGFR 41 at 50-54y | N | GMC (2015-2019): Specimen suboptimal for evaluation. Ultrastructural glomerular features suggesting hereditary nephritis. | Irregular GBM with nonspecific variations in thickness.<br>Global podocyte foot process effacement. | No | Irregular | NA | Absent | NA |
| study_57778 p.Leu1655Arg | 15-19y, M | NA | ESKD | HL | GMC (1990-1994): No biopsy report available. Nephrology note - renal biopsy results reported to patient were post-strep glomerulonephritis. | NA | NA | NA | NA | NA | NA |
| study_78032 c.546+2dup | 35-39y, M | NA | ESKD | N | Unknown (1995-1999): Hereditary Nephritis (Nephrology note) No biopsy report available. | NA | NA | NA | NA | NA | NA |
| study_109651 p.Gln1481Ter | 10-14y, M | G1A3, Y | ESKD | HL | GMC (2000-2004): Alport glomerulopathy, FSGS | Widespread irregularity of GBM with alternating thinning and thickening and foci of splitting and fragmentation of lamina densa. Focal effacement of foot processes. | Y GBM splitting | Y | NA | Mild | NA |
| study_112266<br>c.438+2T>G | 15-19y,<br>M | NA | ESKD | HL | Unknown (2000-2004):<br>hereditary nephritis,<br>questionable<br>membranoproliferative<br>glomerulonephritis | NA | NA | NA | NA | NA | NA |
| study_121508<br>p.Gly624Asp | 45-49y,<br>M | G3bA3, Y | eGFR 14<br>at 55-59y | N | GMC (2010-2014):<br>Suboptimal specimen for a<br>definitive diagnosis. | Focal podocyte foot process<br>effacement<br>Mesangial sclerosis, and some<br>electron-dense material. (1 glom) | No | NR | NA | Mild-<br>moderat<br>e | NA |
| study_88030<br>p.Gly624Asp | 60-64y,<br>M | G3aA3, Y | eGFR 20<br>at 80-84y | HL | Hershey Medical Center<br>(2000-2004): FSGS<br>(Nephrology note) No<br>biopsy report available. | NA | NA | NA | NA | NA | NA |
| **Females** |  |  |  |  |  |  |  |  |  |  |  |
| study_83056,<br>lle355TyrfsTer<br>56 | 5-9y<br>(2<sup>nd</sup><br>biopsy)<br>, F | G1A3, Y<br>(2<sup>nd</sup><br>biopsy) | eGFR 125<br>at 20-24y | N | GMC (2005-2009): IgM<br>nephropathy, FSGS<br>CHOP (2000-2004):<br>insufficient sample, thin<br>basement membrane<br>disease | GMC (2005-2009): GBM segmentally<br>thickened w/segmental sclerotic<br>features. Segment fusion of foot<br>processes.<br>CHOP (2000-2004): GBM focally appear<br>thin (approx. 200nm) but no diffuse<br>thinning. GBM, patchy effacement of<br>epithelial foot processes | GMC (2005-2009):<br>N<br>CHOP (2000-2004): GBM<br>appear focally thin | Focally<br>thin on<br>initial<br>biopsy |  | Minimal | NR |
| study_5080<br>c.546+2dup | 5-9y, F | G1A1, Y | eGFR 94<br>at 20-24y | HL | GMC (2005-2009): No light<br>microscopic or<br>immunofluorescence<br>abnormalities. | NA | NA | NA | NA | Absent | Absen<br>t |
| study_15744<br>p.Gly624Asp | 10-14y,<br>F | NA | eGFR 97<br>at 35-39y | N | Unknown (1990-1994): No<br>biopsy report available.<br>Nephrology note - renal<br>biopsy obtained in<br>Philadelphia - Alport<br>Syndrome diagnosis. No<br>additional details available. | NA | NA | NA | NA | NA | NA |
| study_155572<br>p.Gly624Asp | 35-39y,<br>F | G1A2, Y | eGFR 108<br>at 45-49y | N | GMC (2015-2019): Focal<br>segmental proliferative<br>glomerulonephritis with<br>complement deposits,<br>severe arteriosclerosis,<br>minimal interstitial fibrosis<br>(5%). | Segmental podocyte swelling and<br>effacement; few small associated<br>subepithelial and subendothelial dense<br>deposits and endocapillary<br>proliferation with membrane<br>remodeling. | No | No | NA | Minimal | NA |
| study_167136<br>p.Gly624Asp | Age<br>NA, F | NA | eGFR 76<br>at 50-54y | N | Unknown: No biopsy report<br>available. PCP notes had<br>renal biopsy but provides no<br>other information. | NA | NA | NA | NA | NA | NA |
| study_77063<br>p.Gly624Asp | 40-44y,<br>F | G1A3, Y | eGFR 73<br>at 60-64y | N | GMC (2000-2004):<br>Consistent with an early<br>stage of focal segmental<br>glomerulosclerosis with no<br>significant interstitial<br>fibrosis. | NA | NA | NA | NA | Absent | NA |

Sample/Patient IDs used in Table 4 are unknown to anyone outside the research group. Ages and years are given as a range to preserve anonymity.
This table does not use any USRDS data.
Abbreviations: CKD (chronic kidney disease), : HL (hearing loss), NA (not available), FSGS (focal segmental glomerulosclerosis), GBM (glomerular basement membrane), GS (glomerulosclerosis), EM (electronic microscopy), PCP (primary care provider), IFTA (interstitial fibrosis and tubular atrophy), GMC (Geisinger Medical Center); CKD staging, ACR, PCR, 24 hour urine protein obtained +/- 1 year of biopsy date; CKD-EPI Creatinine Equation (2021) to estimate GFR for adults, Bedside Schwartz equation (2009) used to estimate GFR for pediatrics

### Receipt of Recommended Care for AS

Only 40 (47%) of *COL4A5* patients had ever had ACR testing completed though 82 (96%) had urinalysis data. Overall, only 27 (32%) of *COL4A5* patients were receiving RAAS inhibitors. Among the 37 individuals with ACR ≥30 mg/g or 1+ protein twice on dipstick (and without ESKD), only 18 (49%) were receiving RAAS inhibitors.

## Discussion

In this study using an unselected cohort, we provide more representative quantification of risks associated with X-linked AS, by sex and genotype. Risks of ESKD was highest among males with P/LP variants (44%), intermediate for males with p.Gly624Asp (15%) and females with P/LP variants (10%), and not significantly increased for females with p.Gly624Asp (3%). Overall, we show a wider spectrum of disease severity in both men and women with *COL4A5* variants than described in earlier studies that focused on families known to be afflicted by the disease. For example, prior studies suggested males with X-linked AS having a 100% chance of progression to ESKD^10^. In our cohort, 80% of *COL4A5 P/LP* males and 33% of *COL4A5* p.Gly624Asp males were categorized as having KDIGO high-risk category or greater, and there were 2 men above the age of 70 years who had eGFR ≥60 ml/min/1.73m^2^. This finding of more variable phenotypic severity has important implications in terms of providing more accurate information for families. The broader phenotypic spectrum using a genotype-first approach in an unselected cohort is consistent with other similar genotype-based studies for conditions ranging from autosomal dominant polycystic kidney disease to monogenic diabetes.^5,23,24^ We also uncovered gaps in care related to AS. Despite a median follow up time of approximately 14 years, allowing ample opportunity for various forms of testing, less than half of individuals had albuminuria screening completed and very few individuals with *COL4A5* had a diagnosis of AS.

With our genotype-based approach, we are also able to study genotype-phenotype correlations in a less biased fashion than prior studies. Prior studies have suggested that nonsense/frameshift variants confer the worst prognosis, followed by splice variants, and then the mildest phenotypes for missense and in-frame deletion variants.^6,20^ In our study, we did not observe this genotype-phenotype correlation with similar observed risk for missense variants and PTVs. However, numbers were limited for these analyses, and it is possible that lumping variants into various groups may not reflect differential severity of individual variants within each group. Beyond p.Gly624Asp, which was associated with better outcomes for both males and females, we were unable to make firm conclusions about individual variants. Verifying genotype-phenotype correlations will require larger, additional cohort studies with unselected populations.

Our study and others have shown that p.Gly624Asp, thought to have originated in Central and Eastern Europe due to a founder effect between the 12^th^ and 13^th^ century, confers a milder form of disease and clinical course which may be explained by the location of this variant within the type IV collagen gene^25,26^. The p.Gly624Asp variant is found adjacent to a non-collagenous interruption within the collagenous domain. It has been hypothesized that since p.Gly624Asp occurs adjacent to such an interruption, it likely increases the flexibility of an already flexible region, explaining why it results in a milder form of disease, as compared to variants inducing flexibility in the more rigid parts of the collagenous domain^26^. However, despite being associated with a milder presentation, males with p.Gly624Asp variants still had a 5-fold increased risk of developing eGFR<30 ml/min/1.73m^2^, and 15% had ESKD.

Females are often considered by clinicians as simply being “carriers” of X-linked AS, and we found documentation of this in our chart review of this cohort. Although the manifestations of disease are on average less severe as compared to males, significant kidney risks are still present. An earlier study of families with X-linked AS by Jais et al. had reported a 30% risk of ESKD in females with *COL4A5* by the age of 60, with higher risk in those with concurrent proteinuria and hearing loss^11^. In our study, females with P/LP variants tended to be at increased risk of ESKD with 10% developing ESKD by the age of 65y, likely due to the unselected nature of our cohort. Regardless, the diagnosis of AS in women has important implications for affected women as well as for reproductive decision making and family planning.

Prior research by Gross et al. has demonstrated a role for RAAS inhibitors in delaying onset of renal failure and improving life expectancy in AS^27^. Their study largely focused on AS in males with sibling pairs showing a significantly delayed onset of renal failure in the younger sibling thought to be due to earlier diagnosis and initiation of medical therapy. This delayed onset of renal failure with RAAS blockade was similarly seen in a study examining females with AS^28^. Even in males and females with hypomorphic Gly624Asp variants, RAAS inhibitors have been shown to be associated with reduced risk of end stage renal failure in an observational study by Boeckhaus et al^29^. Despite these studies, only 49% of patients were on RAAS inhibitors even when albuminuria was detected on laboratory studies, emphasizing the importance of early genetic testing and screening to identify such patients. This finding raises the question of whether or not diagnosing these individuals with AS would result in better risk awareness leading to improved adherence to medical therapy, and potentially better patient outcomes.

The major strength of our study is the utilization of an unselected health population and exome sequencing data to inform our diagnosis of patients with X-linked AS, allowing for an in-depth exploration of the phenotypic spectrum of disease. Additionally, due to the availability of extensive longitudinal EHR data, we were able to examine a wide breadth of different phenotypic outcomes. However, EHR data was limited for traits such as ophthalmologic and audiometry data. Another limitation of our study is that all patients with *COL4A5* in our cohort were of European ancestry. In conclusion, we were able to show in a large, unselected cohort, the wide spectrum of phenotypic presentation of males and females with *COL4A5* variants. X-linked AS was underdiagnosed in both males and females with opportunities to improve monitoring of disease progression and better use of guideline-recommended RAAS inhibitors. Further studies are needed to determine whether early genetic diagnosis can improve outcomes in AS.

## Acknowledgements

We are grateful to the many patients who contributed to the MyCode Community Health Initiative by providing genomic and electronic health information. We thank the Regeneron Genetics Center for providing funding for patient enrollment and exome sequencing for the DiscovEHR study, and we would like to acknowledge the Geisinger-Regeneron DiscovEHR Collaboration for the genotypic and phenotypic data. This work was supported by NIGMS grant GM111913 to T.M.

We thank the USRDS for their support. The data reported here have been supplied by the United States Renal Data System (USRDS). The interpretation and reporting of these data are the responsibility of the author(s) and in no way should be seen as an official policy or interpretation of the U.S. government.

## Data availability

The data supporting the findings of this study are available within the article and its Supplementary Data files. Additional information for reproducing the results described in the article is available upon reasonable request and subject to a data use agreement.

## Author contributions

Study design (ARC, HLK), data collection (MZ, KS, KR, BM, IDB, TM), analysis (ARC), interpretation of results (MZ, IDB, TM, ARC), drafted manuscript (MZ, ARC), revised critically for important intellectual content (all authors). All authors have approved the version to be published.

