## Supplemental Materials for "Genotype-first analysis in an unselected health system-based population reveals variable phenotypic severity of *COL4A5* variants"

**Supplemental Methods.**
**Supplemental Table 1. Coding definitions for comorbidities**

**Supplemental Table 2. Modified KDIGO Risk Categories**

**Supplemental Table 3. Firth Logistic Regression Analyses: Phenotypic Features of COL4A5 P/LP X-linked Alport Syndrome by Sex and Genotype**

**Supplemental Table 4. Characteristics by variants**

**Supplemental Table 5. Event Rates and Hazard Ratios for ESKD by Additional Variant Groups in Males and Females**

**Supplemental Table 6. Event Rates and Hazard Ratios for ESKD by Genotype in Males and Females, accounting for competing risk of death**

**Supplemental Table 7. Individuals with a *COL4A5* P/LP variant and an additional rare variant in *COL4A3/4/5***

**Supplemental Table 1. Coding definitions for comorbidities**

| Comorbidities | Codes |
|---|---|
| Hematuria | ICD-10: R31.0, R31.1, R31.21, R31.29, R31.9; ICD-9: 599.7* |
| FSGS | ICD-10: N03.1, N04.1, N05.1, N06.1; ICD-9: 581.1 |
| Alport syndrome | ICD-10: Q87.81 |
| Bilateral sensorineural hearing loss | ICD-10: H90.3; ICD-9: 389.11, 389.12, 389.14, 389.18 |
| Hearing loss | ICD10 H90.*; ICD9 389.* |
| ESKD | Dialysis ICD codes 39.27, 39.42, 39.53, 39.54, 585.6, V45.11, V45.12, V56.1, V56.2, V56.31, V56.32, V56.8, V45.1, N18.6, Z91.15, N18.5+Z99.2<br>Transplant ICD codes 00.91, 00.92, 00.93, 55.53, 55.69, V42.0, 0TY****, Z94.0 |
| Hypertension | ICD-9: 401 - 405<br>ICD-10: I10 - I16 |
| Diabetes | ICD-9: 250*<br>ICD-10: E10, E11, E13 |

**Supplemental Table 2. Modified KDIGO Risk Categories**

| | ACR, mg/g | | |
|---|---|---|---|
| **eGFR** | **<30** | **30-299** | **300+** |
| **60+ without hematuria** | No CKD | Moderately increased risk | High risk |
| **60+ with hematuria** | Hematuria alone | Moderately increased risk | High risk |
| **45-59** | Moderately increased risk | High risk | Very high risk |
| **30-44** | High risk | Very high risk | Very high risk |
| **15-29** | Very high risk | Very high risk | Extremely high risk |
| **<15** | Extremely high risk | Extremely high risk | Extremely high risk |

**Supplemental Table 3. Firth Logistic Regression Analyses: Phenotypic Features of COL4A5 P/LP X-linked Alport Syndrome by Sex and Genotype**

| | Male | | Female | |
|---|---|---|---|---|
| | **P/LP variants (other than Gly624Asp) OR (95%)** | **Gly624Asp OR (95%)** | **P/LP variants (other than Gly624Asp) OR (95%)** | **Gly624Asp OR (95%)** |
| **Any AS phenotypic feature below** | 20.76 (3.60, 119.70)** | 5.37 (1.44, 20.11)* | 4.23 (1.23, 14.56)* | 2.33 (0.99, 5.47) |
| ***ICD diagnosis-based*** | | | | |
| **ESKD ICD** | 40.53 (6.00, 274.00)*** | 3.51 (0.61, 20.04) | 9.95 (1.23, 80.78)* | 2.59 (0.37, 18.33) |
| **Bilateral sensorineural hearing loss ICD** | 18.18 (3.53, 93.68)** | 4.68 (0.45, 48.79) | 1.60 (0.55, 4.66) | 3.45 (0.88, 13.52) |
| **Hearing loss ICD** | 5.00 (1.55, 16.09)** | 10.89 (2.01, 59.14)** | 1.43 (0.32, 6.40) | 1.71 (0.61, 4.81) |
| ***Lab-based diagnoses*** | | | | |
| **Trace blood or greater on UA*** | 4.24 (1.26, 14.29)* | 8.86 (2.20, 35.62)** | 6.26 (1.94, 20.26)** | 2.95 (1.28, 6.82)* |
| **1+ protein twice on UA** | 11.43 (2.96, 44.18)*** | 1.73 (0.46, 6.46) | 4.38 (1.49, 12.94)** | 1.94 (0.82, 4.55) |
| **2+ protein twice on UA** | 11.17 (3.04, 41.05)*** | 2.52 (0.65, 9.84) | 7.29 (2.31, 22.98)** | 3.24 (1.19, 8.84)* |
| **ACR ≥30 mg/g** | 4.65 (0.94, 23.08) | 16.20 (1.64, 159.68)* | 84.64 (4.25, 1683.74)** | 3.85 (1.06, 13.99)* |
| **ACR ≥300 mg/g** | 25.62 (1.21, 543.78)* | 22.14 (0.86, 571.28) | 48.27 (2.26, 1029.82)* | 1.12 (0.16, 7.94) |
| **eGFR<60** | 7.90 (2.39, 26.15)** | 1.77 (0.44, 7.14) | 3.42 (1.19, 9.78)* | 1.42 (0.57, 3.54) |
| **eGFR<30** | 40.53 (6.00, 274.99)*** | 5.38 (1.06, 27.23)* | 5.58 (0.89, 34.95) | 2.74 (0.58, 12.90) |

Firth logistic regression models, only including one individual per family. For kidney transplant patients, eGFR imputed as 5 ml/min/1.73m$^2$
This table does not use USRDS data.
*P<0.05, **P<0.01, ***P<0.001

### Supplemental Table 4. Characteristics by variants

| Variant | ClinVar | REVEL score or Splice AI for splice variants | Gnomad NFE AF | Sex | Age | Hematuria ICD code | Bilateral SNHL ICD code | Hearing loss ICD code | eGFR <60 | eGFR< 30 | ESKD* | Hematuria on dipstick | Proteinuria 1+ on dipstick twice | ACR ≥30 mg/g |
|---|---|---|---|---|---|---|---|---|---|---|---|---|---|---|
| NM_033380.3:c.231+1G>A (n=2 in 1 family) | P (1 star) | SpliceAI: AG 0, AL 0, DG 0.03, DL 0.96 | Absent | Female: 2 | 77.0 (25.5) | 1 (50) | 0 | 0 | 1 (50) | 0 | 0 | 1 (50) | 0 | NA |
| NM_033380.3:c.3017-2A>G (n=3 in 1 family) | P/LP; 2 star | SpliceAI: AG 0.87, AL 0.99, DG 0, DL 0 | Absent | **Male: 2**<br>Female: 1 | **44.0 (48.1)**<br>45-49 | **1 (50)**<br>0 | **0**<br>0 | **1 (50)**<br>0 | **0**<br>0 | **0**<br>0 | **0**<br>0 | **0**<br>0 | **1 (50)**<br>0 | **NA**<br>NA |
| NM_033380.3:c.3553+1G>A (n=1 in 1 family) | NA | SpliceAI: AG 0, AL 0, DL 0.99, DG 0.86 | Absent | Female: 1 | 80-84 | 0 | 0 | 0 | 1 (100) | 0 | 0 | 1 (100) | 0 | 1/1 (100) |
| NM_033380.3:c.438+2T>G (n=2 in 2 families) | NA | SpliceAI: AG 0.03, AL 0.20, DG 0, DL 0.03 | Absent | **Male: 1**<br>Female: 1 | **45-49**<br>60-64 | **0**<br>0 | **1 (100)**<br>0 | **1 (100)**<br>0 | **1 (100)**<br>1 (100) | **1 (100)**<br>1 (100) | **1 (100)**<br>1 (100) | **1 (100)**<br>1 (100) | **1 (100)**<br>0 | **0/1**<br>NA |
| NM_033380.3:c.546+2dup (n=2 in 1 family) | P; 2 stars | SpliceAI: AG 0, AL 0.34, DG 0.01, DL 0.42 | Absent | **Male: 1**<br>Female: 1 | **60-64**<br>20-24 | **0**<br>0 | **0**<br>0 | **0**<br>1 (100) | **1 (100)**<br>0 | **1 (100)**<br>0 | **1 (100)**<br>0 | **0**<br>1 (100) | **0**<br>1 (100) | **0/1**<br>1 (100) |
| NP_203699.1:p.Arg1680Ter (n=2 in 1 family) | P; 2 stars | NA | 1.19E-06 | **Male: 1**<br>Female: 1 | **20-24**<br>50-54 | **0**<br>0 | **1 (100)**<br>0 | **1 (100)**<br>0 | **0**<br>0 | **0**<br>0 | **0**<br>0 | **1 (100)**<br>1 (100) | **1 (100)**<br>1 (100) | NA |
| NP_203699.1:p.Gln1481Ter (n=1 in 1 family) | P; 1 star | NA | Absent | **Male: 1** | **30-34** | **0** | **1 (100)** | **1 (100)** | **1 (100)** | **1 (100)** | **1 (100)** | **0** | **1 (100)** | **0** |
| NP_203699.1:p.Gly1122AspfsTer30 (n=1 in 1 family) | NA | NA | Absent | Female: 1 | 80-84 | 0 | 1 (100) | 1 (100) | 1 (100) | 1 (100) | 1 (100) | 1 (100) | 0 | 1/1 (100) |
| NP_203699.1:p.Gly1170Ser (n=1 in 1 family) | Conflicting (7P, 1 VUS); 1 star | 0.985 | 1.19E-06 | Female: 1 | 50-54 | 1 (100) | 0 | 1 (100) | 0 | 0 | 0 | 1 (100) | 0 | 1/1 (100) |
| NP_203699.1:p.Gly1244Asp (n=5 in 4 families) | P; 2 stars | 0.947 | Absent | **Male: 3**<br>Female: 2 | **33.7 (22.0)**<br>53.0 (17.0) | **2 (67)**<br>0 | **1 (33)**<br>0 | **1 (33)**<br>0 | **1 (33)**<br>1 (50) | **1 (33)**<br>0 | **1 (33)**<br>0 | **3 (100)**<br>2 (100) | **3 (100)**<br>2 (100) | **3/3 (100)**<br>1/1 (100) |
| NP_203699.1:p.Gly307Ser (n=2 in 2 families) | LP; 2 stars | 0.984 | 2.50E-05 | Female: 2 | 27.0 (4.2) | 0 | 0 | 0 | 0 | 0 | 0 | 1 (50) | 1 (50) | NA |
| NP_203699.1:p.Gly624Asp (n=48 in 38 families) | P/LP; 2 stars | 0.91 | 6.15E-05 | **Male: 13**<br>Female: 35 | **55.7 (17.1)**<br>60.3 (14.8) | **0**<br>4 (11) | **1 (8)**<br>3 (9) | **4 (31)**<br>6 (18) | **3 (23)**<br>7 (20) | **3 (23)**<br>2 (6) | **2 (15)**<br>1 (3) | **9/12 (75)**<br>20/33 (61) | **4/12 (33)**<br>12/34 (35) | **4/5 (80)**<br>8/14 (57) |
| NP_203699.1:p.Ile355TyrfsTer56 (n=1 in 1 family) | NA | NA | Absent | Female: 1 | 20-24 | 0 | 0 | 0 | 0 | 0 | 0 | 1 (100) | 1 (100) | NA |
| NP_203699.1:p.Leu1655Arg (n=8 in 5 families) | P; 2 stars | 0.991 | Absent | **Male: 4**<br>Female: 4 | **54.0 (12.4)** | **0**<br>0 | **1 (25)**<br>1 (25) | **3 (75)**<br>2 (50) | **4 (100)**<br>3 (75) | **3 (75)**<br>1 (25) | **3 (75)**<br>1 (25) | **2 (50)**<br>3 (75) | **3 (75)**<br>4 (100) | **2/3 (67)**<br>3/3 (100) |

| | | | | | | | | | | | | | | |
|---|---|---|---|---|---|---|---|---|---|---|---|---|---|---|
| | | | | | 64.5 (28.1) | | | | | | | | | |
| NP_203699.1:p.Pro1050LeufsTer102 (n=1 in 1 family) | NA | NA | Absent | Female: 1 | 85-89 | 0 | 0 | 0 | 0 | 0 | 0 | NA | NA | NA |
| NP_203699.1:p.Pro765GlnfsTer27 (n=1 in 1 family) | P/LP; 2 stars | NA | Absent | Female: 1 | 70-74 | 0 | 0 | 0 | 1 (100) | 0 | 0 | 1 (100) | 1 (100) | 1 (100) |
| NM_033380.3:c.3791-2A>C (n=1 in 1 family) | NA | AG 0.04, AL 0.21, DG 0, DL 0.06 | Absent | **Male: 1** | **70-74** | **0** | **0** | **0** | **0** | **0** | **0** | **0** | **1 (100)** | **1/1 (100)** |
| NM_033380.3:c.3791-2A>G (n=2 in 2 families) | NA | AG 0.03, AL 0.20, DG 0, DL 0.03 | 1.2E-06 | **Male: 2** | **71 (11.3)** | **0** | **0** | **0** | **1 (50) n3** | **0** | **0** | **0** | **1 (50)** | **1/1 (100)** |

For variants with only 1 individuals, age given as a range to preserve anonymity.
Data from https://gnomad.broadinstitute.org/ were accessed on 3/28/24.
This table does not use USRDS data.
Abbreviations: ACMG (American College of Medical Genomics), NFE (non-Finnish European), P(pathogenic), LP (likely pathogenic), LB (likely benign), VUS (variant of unknown significance), NA (not available), BP (benign prediction), PP (pathogenic prediction), eGFR (estimated glomerular filtration rate), ESKD (end-stage kidney disease), ICD (international classification of diseases)

**Supplemental Table 5. Event Rates and Hazard Ratios for ESKD by Additional Variant Groups in Males and Females**

| | Event Rate (per 1000 person-years) | HR (95% CI) | P value |
|---|---|---|---|
| **Males** | | | |
| Control (n=140) | 0.55 (0.20, 1.47) | Ref | |
| Hypomorphic p.Gly624Asp (n=13) | 2.82 (0.71, 11.30) | 5.43 (0.99, 29.78) | 0.051 |
| Missense or in-frame variant (n=7) | 15.51 (5.82, 41.31) | 30.40 (7.26, 127.41) | <0.001 |
| Nonsense or frameshift (n=2) | 24.39 (3.44, 173.15) | 89.46 (4.13, 1939.98) | 0.004 |
| Splice acceptor/donor (n=7) | 5.31 (1.33, 21.21) | 12.35 (2.08, 73.48) | 0.006 |
| **Females** | | | |
| Control (n=262) | 0.26 (0.10, 0.68) | Ref | |
| Hypomorphic p.Gly624Asp (n=35) | 0.48 (0.07, 3.39) | 2.07 (0.23, 18.60) | 0.518 |
| Missense or in-frame variant (n=9) | 2.22 (0.31, 15.78) | 9.84 (1.06, 91.20) | 0.044 |
| Nonsense or frameshift (n=5) | 0 | - | - |
| Splice acceptor/donor (n=6) | 2.94 (0.41, 20.88) | 11.75 (1.18, 116.94) | 0.036 |

**Supplemental Table 6. Hazard Ratios for ESKD by Genotype in Males and Females, accounting for competing risk of death**

| | SHR (95% CI) | P value |
|---|---|---|
| **Males** | | |
| Control (n=140) | Ref | Ref |
| Hypomorphic p.Gly624Asp (n=13) | 7.33 (1.14, 47.08) | 0.036 |
| P/LP variants (n=16) | 24.78 (6.33, 97.04) | <0.001 |
| **Females** | | |
| Control (n=262) | Ref | Ref |
| Hypomorphic p.Gly624Asp (n=35) | 3.45 (0.32, 37.63) | 0.310 |
| P/LP variants (n=20) | 5.83 (0.40, 84.20) | 0.196 |

### Supplemental Table 7. Individuals with a *COL4A5* P/LP variant and an additional rare variant in *COL4A3/4/5*

| Study ID, sex | *COL4A5* P/LP variant | Additional heterozygous rare variant(s) in *COL4A3/4/5* | ClinVar | REVEL Score | Gnomad NFE AF | Phenotype |
|---|---|---|---|---|---|---|
| study_112266, male | NM_033380.3:c.438+2T>G | COL4A4 p.Pro352Leu | Conflicting (3 VUS, 1 benign), 1 star | 0.569 | 0.0002610 | ESKD, dipstick hematuria, bilateral sensorineural hearing loss |
| study_14580, male | NP_203699.1:p.Gly624Asp | COL4A4 p.Pro1241Arg<br>COL4A4 p.Ser1238Ala<br>COL4A4 p.Pro1241Ala | NA<br>NA<br>NA | 0.433<br>0.187<br>0.208 | 8.996e-7<br>absent<br>absent | eGFR 112 at 35-39y, no dipstick proteinuria, no hearing loss ICD |
| study_4155, male | NP_203699.1:p.Gly1244Asp | COL4A5 p.Val918Ile | NA | 0.139 | 0.000004505 | eGFR 126 at 15-19y, dipstick hematuria, ACR 300+, no hearing loss ICD |
| study_52460, male | NP_203699.1:p.Leu1655Arg | COL4A3 p.Val950Ile | Conflicting (3 uncertain, 1 LB), 1 star | 0.332 | 0.0004797 | ESKD, bilateral sensorineural hearing loss ICD, no dipstick hematuria, no dipstick proteinuria |
| study_57778, male | NP_203699.1:p.Leu1655Arg | COL4A3 p.Thr1489Ile | NA | 0.68 | 0.00005000 | ESKD, dipstick hematuria, ACR 300+, no hearing loss ICD |
| study_81593, male | NP_203699.1:p.Gly1244Asp | COL4A5 p.Val918Ile | NA | 0.139 | 0.000004505 | eGFR 97 at 20-24y, dipstick hematuria, ACR 300+, no hearing loss ICD |
| Family A*, study_49551, male | NM_033380.3:c.3017-2A>G | COL4A5 p.Ala430Asp | Benign/Likely Benign, 2 stars | 0.202 | 0.005551 | eGFR 150 at 10-14y, no dipstick hematuria, no hearing loss ICD |
| Family A*, study_77651, male | NM_033380.3:c.3017-2A>G | COL4A5 p.Ala430Asp | Benign/likely benign, 2 stars | 0.202 | 0.005551 | eGFR 77 at 75-59y, no dipstick hematuria, +dipstick proteinuria, no hearing loss ICD |
| Family A*, study_123841, female | NM_033380.37:c.3017-2A>G | COL4A5 p.Ala430Asp | Benign/Likely Benign, 2 stars | 0.202 | 0.005551 | eGFR 92 at 45-49y. No dipstick hematuria, no hearing loss ICD. |
| Family B*, study_140649, female | NP_203699.1:p.Gly1244Asp | COL4A5 p.Val918Ile | NA | 0.139 | 0.000004505 | eGFR 82 at 40-44y, ACR 300+ mg/g, no hearing loss ICD |
| Family B*, study_152355, female | NP_203699.1:p.Gly1244Asp | COL4A5 p.Val918Ile | NA | 0.139 | 0.000004505 | eGFR 46 at 65-59y, dipstick hematuria, 1+ proteinuria on dipstick, no hearing loss ICD |
| study_645, female | NP_203699.1:p.Gly624Asp | COL4A3 p.Glu269Lys<br>COL4A3 p.GLy1277Ser | Benign, 2 stars<br>Conflicting (4 LP, 12 VUS, 1 LB), 1 star | 0.329<br>0.971 | 0.001730<br>0.0003288 | eGFR 90 at 40-49y, dipstick hematuria, no dipstick proteinuria, no hearing loss ICD |
| study_7838, female | NM_033380.3:c.4964T>G | COL4A3 p.Ile1659Val | NA | 0.432 | 0.00002119 | ESKD, hearing loss ICD, ACR 300+, no dipstick hematuria |
| study_78385, female | NP_203699.1:p.Gly624Asp | COL4A4 p.Glu327Gly | NA | 0.428 | 0.00004242 | eGFR 103 at 65-69y, no dipstick hematuria, no dipstick proteinuria, no hearing loss ICD |
| study_132629, female | NP_203699.1:p.Gly1122Asp fsTer30 | COL4A4 p.Ile967Val | Benign, 2 stars | 0.116 | 0.006885 | eGFR 55 at 80-84y, ACR 30-299, bilateral sensorineural hearing loss ICD, no dipstick hematuria |

Data from https://gnomad.broadinstitute.org/ were accessed on 3/28/24.
This table does not use USRDS data and Sample/Patient IDs are not known to anyone outside of the research group.
*Denotes related individuals
Abbreviations: ACMG (American College of Medical Genomics), NFE (non-Finnish European), P(pathogenic), LP (likely pathogenic), LB (likely benign), VUS (variant of unknown significance), NA (not available), BP (benign prediction), PP (pathogenic prediction), eGFR (estimated glomerular filtration rate), ESKD (end-stage kidney disease), ICD (international classification of diseases)
